# Health and Healthcare Variables Associated with Italy’s Excess Mortality during the First Wave of the COVID-19 pandemic: An Ecological Study

**DOI:** 10.1101/2021.01.28.21250669

**Authors:** Alessandra Buja, Matteo Paganini, Riccardo Fusinato, Silvia Cocchio, Manuela Scioni, Vincenzo Rebba, Vincenzo Baldo, Giovanna Boccuzzo

## Abstract

**Background:** Healthcare factors have strongly influenced the propagation of COVID-19. The present study aims to examine whether excess mortality during the first phase of the COVID-19 outbreak in Italy was associated with health, healthcare, demographic, and socioeconomic indicators measured at a provincial level.

**Methods:** The present ecological study concerns the raw number of deaths from Jan. 1 to Apr. 30, 2020 and the mean number of deaths in the same months of 2015 to 2019, per province. Information on socioeconomic factors and healthcare settings were extracted from the most recently updated databases on the ISTAT website. Two multilevel, multivariate models were constructed to test whether excess mortality was associated with the indicators across 107 provinces in Italy.

**Results:** On linear multilevel, multivariate analysis, AIDS mortality rate (p-value <0.05) correlates positively with excess mortality, while a higher density of General Practitioners (number of GPs per 1,000 population) is associated with lower excess mortality (p-value <0.05). After controlling for the diffusion of COVID-19 in each province, the significance of GP density increases (p-value <0.001) and the rate of hospitalization in long-term care wards is positively associated (p-value <0.05) with excess mortality.

**Conclusion:** Some health and healthcare variables are strongly associated with excess mortality caused by COVID-19 in Italy and should be considered to implement mitigation policies and increase healthcare resilience.

## INTRODUCTION

Pandemics are natural disasters that affect global health for prolonged periods [1]. Depending on their impact on healthcare systems, pandemics prompt varying degrees of public health interventions to contain the contagion. The availability of resources – in terms of staff, stuff, and structures – plays a fundamental part in expanding a healthcare system’s capacity to cope with the increasing demand [2]. When healthcare systems are overwhelmed, the level of care delivered to populations gradually declines until the crisis capacity threshold is reached [3]. Beyond that point, severe disruptions in the chain of care affect not only infected patients but also those suffering from chronic illnesses or presenting with acute conditions that need prompt or urgent treatment, leading to a worsening of the population’s health.

Excess mortality is an essential epidemiological indicator for monitoring the capability of healthcare systems. It gives us a comprehensive picture of the death toll attributable to a pandemic. Rapid surveillance systems can effectively track both deaths due directly to the disease and indirect fatalities owing to the impact of a pandemic on healthcare systems [4]. The result is a better mortality estimate, less affected by errors in the certification of the cause of death, that enables before-after comparisons within the same country [5].

As seen previously with the influenza pandemic in 1918 [6], COVID-19 has brought every country to the brink of their healthcare systems’ failure. The first wave of the disease dramatically increased countries’ all-cause mortality rates. Italy registered over 45,000 total deaths in March and April 2020, corresponding to an excess mortality due to any cause of 50% in March and 36% in April compared with the average deaths in the same months of the years from 2015 to 2019 [7].

There were marked differences between regions and provinces, with most excess deaths occurring in the north of Italy. Strict nationwide lockdown policies, which included restrictions on movement between regions from the beginning of March until early May, probably helped contain the infection and the consequent excess mortality in the country’s central and southern regions [8]. The pandemic’s impact on different countries and regions varied [9], but the spread of COVID-19 followed much the same trend in 24 European nations early in the first wave [10]. Social, environmental, and economic factors have strongly influenced the transmission of the disease (especially in the complex scenario of the European continent [9,11]), with a far from a negligible impact on cumulative, all-causes excess deaths.

The present study aims to identify any health, healthcare, environmental, demographic, and socioeconomic indicators associated with the province-level excess mortality during the first wave of the COVID-19 outbreak in Italy. This might reveal underlying “predisposing” factors to bear in mind to implement effective policies to mitigate the effects of a pandemic.

## MATERIALS AND METHODS

### Context

The National Health Service (NHS) in Italy is a regionally-based healthcare system. It provides universal coverage free of charge at the point of service. The country’s central government ensures compliance with the general objectives and fundamental principles of the NHS, defines a national statutory benefits package of “essential levels of care” for the population, and allocates national funds to the regional authorities. The regional governments (of 19 regions plus one region divided into two autonomous provinces, totaling 20 regions) are responsible for organizing and delivering healthcare services through a network of local health authorities. The latter have catchment areas that are often defined by the provinces’ boundaries (institutional entities that group together several municipalities in a given region. Depending on the region, public funds are allocated by local health authorities to public hospitals and accredited private clinics.

### Study design and data sources

The present ecological study includes mortality data for each Italian province. Data on mortality were obtained from the Italian Statistics Institute (ISTAT) demographic database [12] as the raw number of deaths from Jan. 1 to Apr. 30, 2020, and the mean number of deaths during the same months for the years 2015 to 2019.

Information on the provinces’ socioeconomic, demographic, health, and healthcare variables was extracted from the most recently updated databases available on the Italian Ministry of Health’s and the ISTAT’s websites [13], and the ISTAT’s “Health for All” database [14].

### Statistical analysis

The excess all-cause mortality was calculated for each of Italy’s 107 provinces as a percentage. Given the total number of deaths in January-April 2020 for each province (D_1_) and the mean number of deaths during the same period for the years 2015 to 2019 (D_0_), the excess mortality for the i-th province was computed as follows:

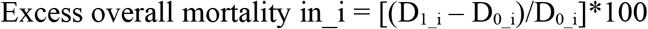

First, a univariate linear regression model was adapted to establish which indicators at provincial level were significantly associated (p-value<0.05 in the univariate model) with the excess mortality. The 70 variables included in the analysis are detailed in Appendix 1. A Pearson’s correlation coefficient was then calculated for each pair of indicators obtained in the previous step to identify those correlating with the smallest number of other indicators.

Six variables regarding healthcare, health, and socioeconomic indicators were selected:

- acquired immunodeficiency syndrome (AIDS) mortality rate: number of deaths due to AIDS per 1,000 population;
- long-term care hospitalization rate: number of hospital admissions in long-term care wards per 1,000 population;
- gross domestic product per capita based on purchasing power parity (GDP);
- average density of general practitioners (GPs): number of GPs per 1,000 population;
- average density of hospital physicians: number of hospital physicians per 10,000 population;
- province’s COVID-19 transmission factor: a measure of the rate at which COVID-19 spread in each province.

This last variable, the transmission factor, measures the trend of contagion as the relative increase in the number of individuals infected between two time points, T0 and T1, and it was calculated under the hypothesis of exponential growth [11]. The data were drawn from the COVID-19 Italian Civil Protection Department’s official database, which was updated daily [15]. The date T0 was set as the day when a given province reached at least 10 total cases, and the date T1 was 30 days later.

Two multivariate models were developed. Model 1 is a multilevel, multivariate linear model including the first five province-related indicators on the first level and the region as a grouping variable on the second to consider the different rates of transmission of the infection in other areas. Model 2 is an adjusted linear multivariate model in which we add the province’s COVID-19 transmission factor as a covariate. Since this indicator measures the rate of COVID-19 spread in each province, we can expect it to capture most of the variability in the outcome, enabling us to better understand the role of the other covariates as excess mortality risk factors.

Quantile regression was also conducted, and the parameter estimates of each covariate variable were plotted at different quantiles of the outcome variable (excess mortality). The data analysis was performed with SAS Studio statistical software (ver. 5.2, SAS Institute Inc., Cary, NC).

## RESULTS

In the period considered, excess mortality ranged from -10.99% (for L’Aquila, a province in the central Abruzzo region) to 164.20% (for Bergamo, in the northern Lombardy region), with a mean value of 15.02%, and a median of 4.43% across Italy as a whole. The five provinces with the lowest excess mortality are in Southern Italy, while the five with the highest excess mortality are all in Northern Italy: four in Lombardy, and one (Piacenza) in the Emilia-Romagna region on Lombardy’s southern border (see Table 1). Table 2 shows the descriptive statistics for the six indicators selected.

**Table 1.**
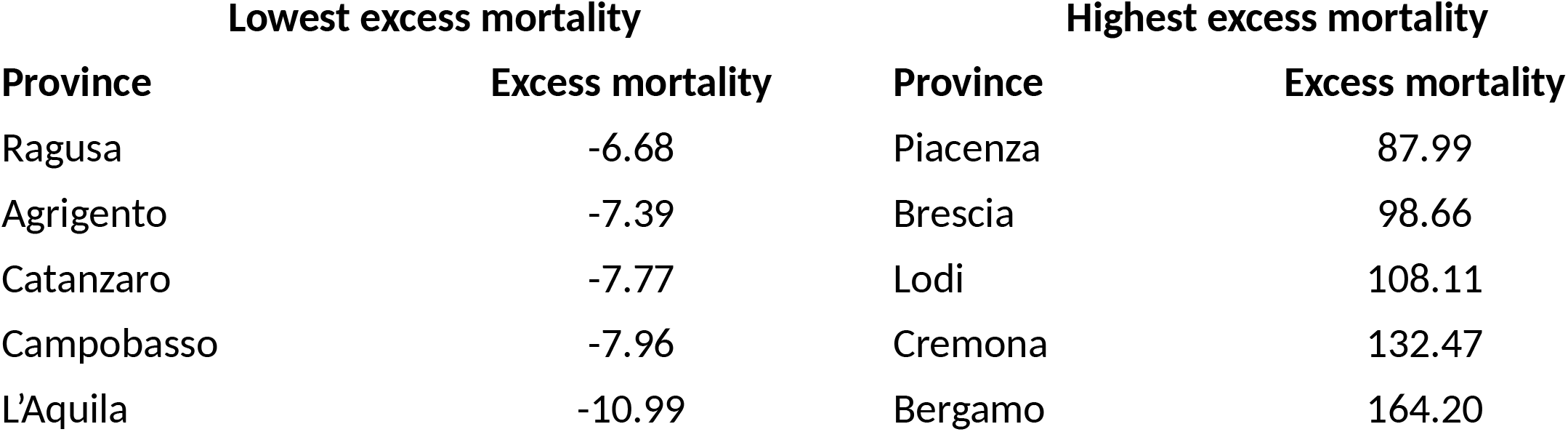
Provinces with the five highest and five lowest excess mortality rates (percentages)

| Lowest excess mortality |  | Highest excess mortality |  |
| --- | --- | --- | --- |
| Province | Excess mortality | Province | Excess mortality |
| Ragusa | -6.68 | Piacenza | 87.99 |
| Agrigento | -7.39 | Brescia | 98.66 |
| Catanzaro | -7.77 | Lodi | 108.11 |
| Campobasso | -7.96 | Cremona | 132.47 |
| L'Aquila | -10.99 | Bergamo | 164.20 |

**Table 2.** Descriptive statistics for the explanatory variables included in the models

| Variables | Minimum | Maximum | Mean | Median | Standard deviation | IQR |
| --- | --- | --- | --- | --- | --- | --- |
| Province's transmission factor | 0.05 | 0.26 | 0.14 | 0.14 | 0.04 | 0.06 |
| AIDS mortality rate | 0 | 0.27 | 0.07 | 0.06 | 0.05 | 0.07 |
| Gross domestic product per capita | 15000 | 53400 | 26009 | 26000 | 7271 | 12100 |
| Long-term hospitalization rate | 0 | 34.41 | 7.11 | 6.34 | 5.09 | 6.38 |
| Number of hospital physicians per 10,000 population | 8.32 | 33.61 | 19.74 | 19.53 | 5.19 | 6.88 |
| Number of GPs per 1,000 population | 0.7 | 1.3 | 0.94 | 0.9 | 0.12 | 0.1 |

Figure 1 shows the trend of the indicators estimated for all excess mortality quantiles.

**Figure 1.**
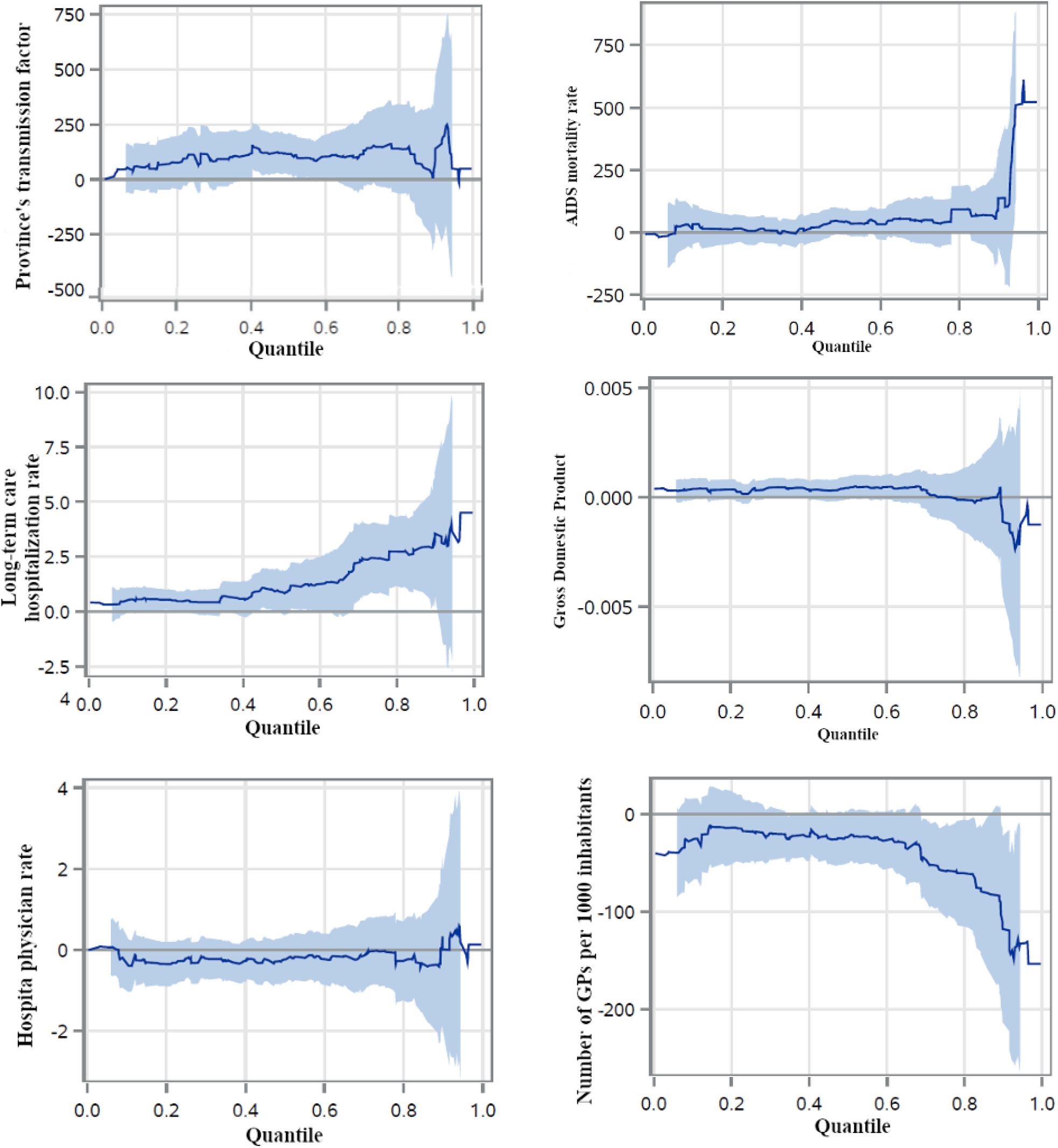
Results of quantile regression: variation in estimated parameters at different quantiles of excess mortality (with 95% confidence intervals)

Table 3 shows the estimates for the unadjusted associations and the results of the two multivariate models. In Model 1 (linear multilevel, multivariate analysis with the first five province-related indicators), the AIDS mortality rate (p-value <0.05) is positively associated with excess mortality, and the long-term care hospitalization rate is statistically significant at p-value ∼0.10. The density of GPs is negatively associated with the outcome variable (p-value <0.05). After controlling for the rate at which COVID-19 spread in each province, the linear multivariate Model 2 suggests a significant positive association (p-value <0.05) between excess mortality and a province’s transmission factor, AIDS mortality rate, and long-term care hospitalization rate. The density of GPs is significantly and negatively associated with excess mortality (p-value <0.001).

**Table 3.** Regression coefficients for variables explaining excess mortality

| **Variable** | **Unadjusted**<br>**coefficients (p-**<br>**value)** | **Multivariate Model**<br>**1 coefficients (p-**<br>**value)** | **Multivariate Model 2**<br>**(adjusting for provinces'**<br>**transmission factor)**<br>**(p-value)** |
| --- | --- | --- | --- |
| AIDS mortality rate | **187.76 (0.001)** | **63.044 (0.043)** | **97.192 (0.037)---** |
| Long-term care<br>hospitalization rate | **2.50 (<0.001)** | 0.774 (0.106) | **1.13 (0.023)** |
| GDP | **0.002 (<0.001)** | 0.001 (0.251) | ---0.000 (0.876) |
| No. of hospital<br>physicians per 10,000<br>population | **0.49 (0.037)** | --0.340 (0.464) | -0.017 (0.973)-- |
| No. of GPs per 1,000<br>population | **-130.573 (<0.001)** | **-71.499 (0.005)---** | **-82.612 (0.0008)---** |
| Province's<br>transmission factor | **361.72 (<0.001)** | --- | **173.96 (0.030)** |

It is worth noting that, with both multivariate analyses (Models 1 and 2), GDP per capita loses its significance as a covariate positively correlated with excess mortality. This may stem from the fact that socioeconomic factors (capturing economic and social interactions and transportation flows) tend to be more significantly related to COVID-19 incidence than to excess mortality [11,16]. Simultaneously, health and healthcare variables become more important in explaining the excess mortality directly and indirectly caused by COVID-19 in the various provinces.

## DISCUSSION

Since the outbreak of COVID-19 in Europe, strict public health measures have gradually been introduced to contain the contagion. Decision-makers have relied mostly on daily updates regarding new cases of COVID-19, COVID-19 mortality, the numbers of those infected being admitted to general wards and intensive care units, and the number of patients who have recovered. The under-reporting of cases, unfortunately, hindered the proper interpretation of these data. On the other hand, excess mortality can be more useful in describing pandemic trends in relation to other variables affecting local and national health systems [17].

This ecological study found that some health and healthcare variables were associated with excess mortality at a provincial level during the first wave of the COVID-19 outbreak in Italy (January-April 2020). Our analysis confirmed that most excess deaths occurred in northern provinces in or adjacent to Lombardy, the region mostly overwhelmed by the pandemic [8].

Judging from our Model 1 (considering the first five province-related independent variables), the excess mortality caused directly and indirectly by COVID-19 across Italy’s provinces correlated positively with the AIDS mortality rate and negatively with a higher density of GPs. Adding the province’s COVID-19 transmission factor, Model 2 confirmed the significance of the AIDS mortality rate, while both the density of GPs and the long-term care hospitalization rate increased in significance as variables explaining excess mortality.

Our finding of a significant positive association between excess mortality due to COVID-19 and AIDS mortality in recent years is interesting, especially in light of these two infectious diseases’ different transmission routes. AIDS prevention and treatment are among the most important public health achievements of recent decades. They are also strongly influenced by socio-cultural factors and by individual behavior and attitudes [18]. The positive association identified between excess mortality due to COVID-19 and AIDS mortality could result from a complex interaction between the public health system, socio-cultural environments, and individual lifestyles and behavior (e.g., positive prevention practices and adherence to antiviral medication among individuals with HIV). According to this interpretation, local communities with higher AIDS mortality rates, owing to a relatively lower propensity for prevention and more risky behavior not adequately countered by effective public health services, could also have higher excess mortality due to COVID-19. This would show that behavior and attitudes to prevention are important social and individual characteristics associated with how people respond to COVID-19 prevention. This should be borne in mind in future public health efforts to improve adherence to recommendations for controlling the spread of infectious diseases.

A higher density of GPs was significantly associated with a lower COVID-19 excess mortality. This indicator could be seen as a measure of the accessibility to primary care. Its negative association with the outcome variable suggests that provinces with fewer GPs per 1,000 population may have experienced a higher COVID-19 excess mortality because GPs faced an excessive workload at the onset of the pandemic, making them less able to meet their patients’ needs. Primary care has a crucial role in minimizing the burden of COVID-19 cases in hospitals. GPs can help prevent healthcare system’s overload, but they need appropriate engagement and advice to work effectively as gatekeepers to acute care facilities [19]. Primary care services should have response mechanisms and systems in place well before there is widespread community transmission to ensure the continued availability of services and enable a rapid upscaling of services as required. People exposed to COVID-19 and seeking a test or medical advice may have had great difficulty accessing primary care. When people cannot access a primary care provider for routine or COVID-19-related care, they may flood emergency departments and urgent care facilities, contributing to overwhelm the hospital system and potentially increasing the transmission of COVID-19. Besides, patients with complex medical needs (such as multiple chronic conditions, functional limitations, and other health issues) may have been told only to seek care for urgent problems, but their delayed management may have worsened their condition. Several policies could help primary care to build capacity in the short term and mitigate the effects of future crises. For instance, policymakers can help primary care providers develop an adequate telehealth infrastructure and offer them fair reimbursement. A more robust and well-functioning primary care system is now needed more than ever to save lives in an increasingly daunting health crisis [20].

Our Model 2 considered all six province-related indicators as covariates, including the COVID-19 transmission factor (which considers the different rates of COVID-19 transmission across Italy’s provinces). Adding this transmission factor captures most of the variability in excess mortality at a province level. The model nonetheless confirms the significance of AIDS mortality and the density of GPs as explanatory variables. The long-term care hospitalization rate also becomes significantly and positively associated with excess mortality. This result may mean that, after controlling for the rate of COVID-19 transmission across provinces, excess mortality is strongly influenced by the incidence of individuals with a high health risk among the population infected. In other words, a large part of the excess deaths was attributable to the elderlies admitted to long-term care wards. Such patients are typically frailer (owing to pre-existing conditions and chronic diseases, or specific rehabilitation needs) and at substantial risk of dying once infected, as are nursing home residents [21]. This factor seems to have contributed to the excess mortality seen in several European countries during the first wave of COVID-19 when specific measures to control the disease’s spread (personal protective equipment, proper isolation of infected cases, training of personnel on infectious disease management) were in the early stages, and the likelihood of nosocomial transmission of COVID-19 was still high [22]. Moreover, even if they were not infected with COVID-19, patients in long-term care settings had to be abruptly isolated, losing contact with families and caregivers. Such a situation may have led to depression and a decline in their psychological and physical health [23]. Solutions targeting these specific issues and ways to improve the community health training of long-term care personnel and administrators should be investigated in the future. From this perspective, it also appears vital to strengthen primary care services. The provision of intermediate services (community hospitals and rehabilitation facilities), home care services, and telecare solutions in Italy are currently insufficient to cope with the needs of the frail elderly, which often leads to inappropriate admissions to acute care hospitals.

### Limitations

The main limitation of this study lies in the relatively small province sample size, which could lend a low power to the statistical analyses. These findings also only regard the Italian context and may not hold for other countries. Finally, it is worth mentioning that any correlations emerging from ecological studies do not necessarily imply causality between two events; this could only emerge from more extensive studies and with the support of more complex statistical methods.

## CONCLUSIONS

This ecological study indicates that some health and healthcare variables were strongly associated with the excess mortality caused directly and indirectly by the COVID-19 outbreak in Italy during the first wave of the pandemic (January-April 2020). These variables may be seen as drivers behind the spatially uneven spread of COVID-19, with most excess deaths occurring in some parts of northern Italy. They should be taken into account to implement effective mitigation policies and make health systems more resilient to COVID-19 and other crises by reshaping the organization of healthcare resources and services.

Our results suggest that knowing more about different territories’ susceptibility to the diffusion of viruses (which is linked to their propensity for prevention) may help prepare locally-tailored mitigation strategies in countries like Italy, which are characterized by a marked heterogeneity across local communities.

Our findings also provide important input for the design of policies aiming to improve a health system’s resilience by drawing on the Italian NHS’s experience during the first wave of COVID-19. This study shows that the excess mortality due to COVID-19 may be fueled by a limited capacity to respond to primary care and community care services. Therefore, it seems crucial to readjust the balance in the allocation of resources and strengthen these areas of public healthcare.

## Data Availability

Data used in this study are available on the referenced public datasets.

## Acknowledgments

This work was carried out within the framework of the research project “Impact of different healthcare System MOdels and of different COntainment measures on the spread and health outcomes of COVID-19 in Italy and Europe” (MOSSCOV) – University of Padova.

## Authors’ contributions

AB, GB: Conceptualization; MS, VR: Data curation; RF, AB, MS, GB: Formal analysis; AB, VR, VB, GB: Funding acquisition; RF, AB, MS, GB: Investigation; AB, GB: Methodology; AB, GB: Project administration; AB, VR, VB, GB: Resources; AB, GB: Software; AB, VR, VB, GB Supervision; AB, VR, VB, GB: Validation; AB, GB: Visualization; MP, AB; VB, VR: Writing - original draft; MP, AB; VB, VR: Writing - review & editing.

## Data availability

data used in this study are available on the referenced public datasets.

