## Appendix 1 for "Health and Healthcare Variables Associated with Italy’s Excess Mortality during the First Wave of the COVID-19 pandemic: An Ecological Study"

**Appendix 1.** List of variables considered for the analysis by province (70 variables)

| Variable | Type | Reference year | Definition | Source | Outcome of the univariate linear regression model |
|---|---|---|---|---|---|
| **Aging index** | Demographic | 2018 | (65+ Population) / (0-14 Population) * 100 | Health for All | Not significant in the univariate model |
| **Dependency ratio** | Demographic | 2018 | (0-14 Population + 65+ Population) / (15-64 Population) * 100 | Health for All | Not significant in the univariate model |
| **Number of foreign residents** | Demographic | 2019 | | ISTAT | Not significant in the univariate model |
| **Number of residence permits** | Demographic | 2019 | | ISTAT | Not significant in the univariate model |
| **Population density** | Demographic | 2020 | (Population) / (Area) | ISTAT | Not significant in the univariate model |
| **PM10 emissions (in gigagrams)** | Environment | 2015 | | ISPRA | Not significant in the univariate model |
| **AIDS mortality rate** | Health | 2017 | (Deaths due to AIDS) / (Population) * 10,000 | Health for All | Selected |
| **Cardiovascular diseases mortality rate** | Health | 2017 | (Deaths due to cardiovascular disease) / (Population) * 10,000 | Health for All | Not selected because of the high correlation with other selected variables |
| **Hospital discharge rate with diagnosis of diabetes** | Health | 2016 | (Diabetes patients discharged) / (Population) * 100 | Health for All | Not significant in the univariate model |
| **Hospital discharge rate with diagnosis of malignant neoplasm** | Health | 2016 | (Malignant neoplasm patients discharged) / (Population) * 100 | Health for All | Not significant in the univariate model |
| **Infectious disease mortality rate** | Health | 2017 | (Deaths due to infectious disease) / (Population) * | Health for All | Not selected because of the high correlation with other selected variables |

10,000

| Variable | Type | Reference year | Definition | Source | Outcome of the univariate linear regression model |
|---|---|---|---|---|---|
| **Province's transmission factor** | Health | 2020 | | Calculated | Selected |
| **Psychiatric disease mortality rate** | Health | 2017 | (Deaths due to psychiatric disease) / (Population) * 10,000 | Health for All | Not selected because of the high correlation with other selected variables |
| **Trauma mortality rate** | Health | 2017 | (Trauma deaths) / (Population) * 1,000 | Health for All | Not significant in the univariate model |
| **Tuberculosis mortality rate** | Health | 2017 | (Tuberculosis deaths) / (Population) * 1,000 | Health for All | Not selected because of the high correlation with other selected variables |
| **Acute hospital admission rate** | Healthcare activity | 2016 | (Acute patients) / (Population) * 1,000 | Health for All | Not significant in the univariate model |
| **Acute admission rate in public hospitals** | Healthcare activity | 2016 | (Acute patients in public hospitals) / (Population) * 1,000 | Health for All | Not significant in the univariate model |
| **Acute admission rate in private hospitals** | Healthcare activity | 2016 | (Acute patients in private hospitals) / (Population) * 1,000 | Health for All | Not significant in the univariate model |
| **Hospital acute patients discharge rate** | Healthcare activity | 2016 | (Acute patients discharged) / (Population) * 100 | Health for All | Not significant in the univariate model |
| **Hospital admission rate** | Healthcare activity | 2016 | (Patients) / (Population) * 1,000 | Health for All | Not significant in the univariate model |
| **Admission rate in private hospitals** | Healthcare activity | 2016 | (Patients in private hospitals) / (Population) * 1,000 | Health for All | Not significant in the univariate model |
| **Admission rate in public hospitals** | Healthcare activity | 2016 | (Patients in public hospitals) / (Population) | Health for All | Not significant in the univariate model |

*1,000

| Variable | Type | Reference year | Definition | Source | Outcome of the univariate linear regression model |
|---|---|---|---|---|---|
| **Ambulance usage rate in emergency medical service** | Healthcare activity | 2016 | (Number of ambulances) / (Population) * 100,000 | Health for All | Not significant in the univariate model |
| **Hospital discharge rate of patients treated with chemotherapy** | Healthcare activity | 2016 | (Chemotherapy patients discharged) / (Population) * 100 | Health for All | Not selected because of the high correlation with other selected variables |
| **Emergency room access rate per 1000 population** | Healthcare activity | 2016 | (Number of ER accesses) / (Population) * 100 | Health for All | Not significant in the univariate model |
| **Bed occupancy rate** | Healthcare activity | 2016 | (Used beds) / (Available beds) * 100 | Health for All | Not significant in the univariate model |
| **Hospitalization rate in long-term care wards** | Healthcare activity | 2016 | (Patients in long-term care) / (Population) * 1,000 | Health for All | Selected |
| **Hospitalization rate in long-term care wards of private hospitals** | Healthcare activity | 2016 | (Patients in long-term care in private hospitals) / (Population) * 1,000 | Health for All | Not selected because of the high correlation with other selected variables |
| **Hospitalization rate in long-term care wards of public hospitals** | Healthcare activity | 2016 | (Patients in long-term care in public hospitals) / (Population) * 1,000 | Health for All | Not selected because of the high correlation with other selected variables |

| Variable | Type | Reference year | Definition | Source | Outcome of the univariate linear regression model |
|---|---|---|---|---|---|
| **Average length of stay in acute hospitals** | Healthcare activity | 2016 | | Health for All | Not selected because of the high correlation with other selected variables |
| **Average hospital personnel density (No. of hospital workers per 10,000 population)** | Healthcare human resources | 2016 | (Personnel) / (Population) * 10,000 | Health for All | Selected |
| **Average hospital physician density (No. of hospital physicians per 10,000 population)** | Healthcare human resources | 2016 | (Physicians) / (Population) * 10,000 | Health for All | Selected |
| **Average hospital nurse density (No. of hospital nurses per 10,000 population)** | Healthcare human resources | 2016 | (Nurses) / (Population) * 10,000 | Health for All | Not selected because of the high correlation with other selected variables |
| **Number of cardiologists** | Healthcare human resources | 2018 | | Health for All | Not significant in the univariate model |
| **Number of intensive care physicians** | Healthcare human resources | 2018 | | Health for All | Not significant in the univariate model |
| **Number of surgeons** | Healthcare human resources | 2018 | | Health for All | Not significant in the univariate model |

| Variable | Type | Reference year | Definition | Source | Outcome of the univariate linear regression model |
|---|---|---|---|---|---|
| **Nurses-to-beds ratio** | Healthcare human resources | 2016 | (Nurses) / (Beds) * 100 | Health for All | Not selected because of the high correlation with other selected variables |
| **Percentage of private hospital nurses out of total hospital nurses** | Healthcare human resources | 2016 | (Private hospital nurses) / (Nurses) * 100 | Health for All | Not selected because of the high correlation with other selected variables |
| **Percentage of private hospital personnel out of total hospital personnel** | Healthcare human resources | 2016 | (Private hospital personnel) / (Hospital personnel) * 100 | Health for All | Not selected because of the high correlation with other selected variables |
| **Percentage of private hospital physicians out of total hospital physicians** | Healthcare human resources | 2016 | (Private hospital physician) / (Hospital physicians) * 100 | Health for All | Not selected because of the high correlation with other selected variables |
| **Physicians-to-beds ratio** | Healthcare human resources | 2016 | (Physician) / (Beds) | Health for All | Not selected because of the high correlation with other selected variables |
| **Physicians per 1,000 people** | Healthcare human resources | 2018 | (Physicians) / (Population) * 1,000 | Health for All | Not selected because of the high correlation with other selected variables |
| **Personnel per 1,000 people in public healthcare** | Healthcare human resources | 2016 | (Personnel in public health) / (Population) * 1,000 | Health for All | Not selected because of the high correlation with other selected variables |
| **Nurses per 1,000 people in public healthcare** | Healthcare human resources | 2016 | (Nurses in public health) / (Population) * 1,000 | Health for All | Not selected because of the high correlation with other selected variables |

| Variable | Type | Reference year | Definition | Source | Outcome of the univariate linear regression model |
|---|---|---|---|---|---|
| **Physicians per 1,000 people in public healthcare** | Healthcare human resources | 2016 | (Physician in public health) / (Population) * 1,000 | Health for All | Not selected because of the high correlation with other selected variables |
| **Physicians registered with the National Board of Physicians per 1,000 people** | Healthcare human resources | 2018 | (Registered physicians) / (Population) * 1,000 | Health for All | Not significant in the univariate model |
| **General practitioners per 1,000 people** | Healthcare human resources | 2019 | (General practitioners) / (Population) * 1,000 | Sole24Ore Quality of Life Index 2020 (based on Ministry of Health data) | Selected |
| **Number of acute care beds in hospitals** | Healthcare resources | 2016 | | Health for All | Not significant in the univariate model |
| **Number of acute care beds in private hospitals** | Healthcare resources | 2016 | | Health for All | Not significant in the univariate model |
| **Number of acute care beds in public hospitals** | Healthcare resources | 2016 | | Health for All | Not significant in the univariate model |
| **Number of long-term care beds in private hospitals** | Healthcare resources | 2016 | | Health for All | Not selected because of the high correlation with other selected variables |
| **Number of long-term care beds in public hospitals** | Healthcare resources | 2016 | | Health for All | Not selected because of the high correlation with other selected variables |

| Variable | Type | Reference year | Definition | Source | Outcome of the univariate linear regression model |
|---|---|---|---|---|---|
| **Number of long-term care beds in hospitals** | Healthcare resources | 2016 | | Health for All | Not selected because of the high correlation with other selected variables |
| **Number of acute hospital beds** | Healthcare resources | 2016 | | Health for All | Not selected because of the high correlation with other selected variables |
| **Number of acute care beds in private hospitals** | Healthcare resources | 2016 | | Health for All | Not significant in the univariate model |
| **Number of acute beds in public hospitals** | Healthcare resources | 2016 | | Health for All | Not selected because of the high correlation with other selected variables |
| **Percentage of hospitals with intensive care units.** | Healthcare resources | 2016 | (Hospitals with ICU) / (Hospitals) * 100 | Health for All | Not significant in the univariate model |
| **Percentage of private hospital acute care beds on total acute care hospital beds** | Healthcare resources | 2016 | (Private acute care beds) / (Acute care beds) * 100 | Health for All | Not significant in the univariate model |
| **Percentage of private hospital long-term care beds out of total long-term care hospital beds** | Healthcare resources | 2016 | (Private long-term care beds) / (Long-term care beds) * 100 | Health for All | Not significant in the univariate model |

| Variable | Type | Reference year | Definition | Source | Outcome of the univariate linear regression model |
|---|---|---|---|---|---|
| **Percentage of private hospital acute care beds out of total ordinary care hospital beds** | Healthcare resources | 2016 | (Private acute care beds) / (Ordinary beds) * 100 | Health for All | Not significant in the univariate model |
| **General practitioners per 1,000 people** | Healthcare resources | 2020 | General practitioners / (Population) * 1,000 | Sole 24 ore | Selected |
| **Economic activity rate** | Socio-economic | 2019 | (Workforce population / 15+ population) * 100 | Health for All | Not selected because of the high correlation with other selected variables |
| **Employment rate** | Socio-economic | 2019 | (15+ People working) / (15+ Population) | ISTAT | Not selected because of the high correlation with other selected variables |
| **Gross domestic product per capita** | Socio-economic | 2016 | | SISREG | Selected |
| **Number of industrial firms** | Socio-economic | 2018 | | ISTAT | Not significant in the univariate model |
| **Number of workers in the industrial firms** | Socio-economic | 2018 | | ISTAT | Not significant in the univariate model |
| **Poverty index** | Socio-economic | 2018 | (In poverty population) / (Population) * 100 | ISTAT | Not selected because of the high correlation with other selected variables |
| **Local public transport facility availability** | Socio-economic | 2015 | (Buses + Metro) / (population) | ISTAT | Not significant in the univariate model |
| **Local public transport use per capita** | Socio-economic | 2015 | (Annual passengers) / (Population) | ISTAT | Not significant in the univariate model |
| **Unemployment rate** | Socio-economic | 2019 | (35+ Job seekers) / (35+ Workforce population) | ISTAT | Not selected because of the high correlation with other selected variables |
